# Predictive Model of Risk Factors of High Flow Nasal Cannula Using Machine Learning in COVID-19

**DOI:** 10.1101/2022.03.02.22271673

**Authors:** Nobuaki Matsunaga, Keisuke Kamata, Yusuke Asai, Shinya Tsuzuki, Yasuaki Sakamoto, Shinpei Ijichi, Takayuki Akiyama, Jiefu Yu, Gen Yamada, Mari Terada, Setsuko Suzuki, Kumiko Suzuki, Sho Saito, Kayoko Hayakawa, Norio Ohmagari

## Abstract

**Background:** With the rapid increase in the number of COVID-19 patients in Japan, the number of patients receiving oxygen at home has also increased rapidly, and some of these patients have died. An efficient approach to identify high-risk patients with slowly progressing and rapidly worsening COVID-19, and to avoid missing the timing of therapeutic intervention will improve patient prognosis and prevent medical complications.

**Methods:** Patients admitted to medical institutions in Japan from November 14, 2020 to April 11, 2021 and registered in the COVID-19 Registry Japan were included. Risk factors for patients with High Flow Nasal Cannula invasive respiratory management or higher were comprehensively explored using machine learning. Age-specific cohorts were created, and severity prediction was performed for the patient surge period and normal times, respectively.

**Results:** We were able to obtain a model that was able to predict severe disease with a sensitivity of 57% when the specificity was set at 90% for those aged 40-59 years, and with a specificity of 50% and 43% when the sensitivity was set at 90% for those aged 60-79 years and 80 years and older, respectively. We were able to identify lactate dehydrogenase level (LDH) as an important factor in predicting the severity of illness in all age groups.

**Discussion:** Using machine learning, we were able to identify risk factors with high accuracy, and predict the severity of the disease. Using machine learning, we were able to identify risk factors with high accuracy, and predict the severity of the disease. We plan to develop a tool that will be useful in determining the indications for hospitalisation for patients undergoing home care and early hospitalisation.

## Introduction

Since the end of 2019, the new coronavirus infection caused by SARS-CoV-2 (COVID-19) has spread globally from China and become a public health crisis worldwide [1,2]. In particular, the establishment of a medical system to treat the rapid increase in the number of patients with COVID-19 has become an urgent issue; this could be one of the reasons for the excessive mortality [3]. In Japan, all patients with COVID-19 from January 2020 onwards were managed on an inpatient basis [4].

With the increase in patients with COVID-19, the hospitalisation criteria have been gradually increased, and at the discretion of the local governors, since November, patients with moderate– severe illness are considered for hospitalisation [5]. However, in the early 2021, with the rapid increase in the number of patients with COVID-19, it became difficult to examine all patients with hypoxemia in the hospital, and there were patients who received oxygen therapy at home or hotel medical facilities, whereas some died [6].

In the COVID-19 Registry Japan (COVIREGI-JP), which is run and managed by the National Centre for Global Health and Medicine (NCGM), COVID-19 inpatient data is recorded so that evidence related to epidemiology, prevention and treatment can be collected. In September 2021, over 50,000 patients were registered, making it a registry that represents the entire Japan [7]. With regard to COVID-19 in Japan, there have been reports of descriptive studies, and risk scores have been provided to estimate the risk factors for progression to severe disease and death; however, the accuracy of these data needs to be improved. Studies have been conducted using machine learning to extract risk factors from the enormous volume of medical data [8–10]. It was thought that studies using machine learning are needed.

Identifying risk factors is important in patients who require invasive respiratory management rather than adjuvant ventilation therapy (High Flow Nasal Cannula) so that the decision to transport patients to an advanced medical care facility can be made during medical crises. Some reports that examined risk factors for requiring invasive artificial respiratory management and death can be found; however, to our knowledge, there are no reports that examined the risks for requiring invasive respiratory management rather than nasal high flow from the perspective of health resource distribution. In patients with COVID-19 who slowly progress and undergo rapid disease exacerbation, identification of risk factors for progression to severe disease in detail and observation so that timely treatment can be performed are important to improve prognosis and prevent the collapse of the medical system.

Therefore, in this paper, using machine learning, we aimed to comprehensively search for and identify risk factors in patients who underwent invasive respiratory treatment rather than nasal high flow after hospitalisation, and by confirming the criteria for at-home medical care, hotel medical care and inpatient management, we aimed to enable safe and efficient utilisation of medical resources.

## Methods

### Study design and patients

This is an observational study. Medical institutions that participated voluntarily in the COVIREGI-JP enrolled patients. Study co-operators at each institution input data into case report forms electronically and manually. Co-researchers obtained funding from a grant-in-aid for each patient who was enrolled. The patient enrollment criteria were as follows: 1) having a positive SARS-CoV-2 test result and 2) receiving inpatient treatment at a medical institution. Among the inpatients eligible for the study, there were some patients who were not registered in the COVIREGI-JP at the discretion of the principal investigator (e.g., patients who participated in other clinical research, whose data was deemed unsuitable for registration in the COVIREGI-JP and those who refused study participation by opting out). Each hospitalisation was registered for patients with a history of multiple hospitalisations due to COVID-19 and those who satisfied the aforementioned inclusion criteria.

### Data Collection and Case Report Form

The case report form of the International Severe Acute Respiratory and Emerging Infection Consortium was revised to enable the collection of clinical, epidemiological and treatment data of Japan. Research data was collected and managed using Research Electronic Data Capture (REDCap) [11], which is a safe web-based data capture application hosted by the Joint Center for Researchers, Associates and Clinicians, a data centre of the NCGM.

### Datasets

We used the data of patients for whom all primary items, such as demographics, epidemiological characteristics, comorbidities, signs and symptoms, and results of laboratory examination at the time of hospitalisation, outcome at the time of discharge and supportive therapy, were recorded as of July 1, 2021. Parameters indicating ‘unknown’ in the option were not treated as missing data. In the test values, those with clear unit errors were corrected. Obvious outliers were excluded and treated as missing data. Two infectious disease specialists were consulted, and test values that are not usually acquired were not used as factors. As for signs and symptoms and results of laboratory examination, we included only those at the time of admission. Furthermore, the features that were input as text were converted into numerical values. Smoking and drinking habits at admission were recorded as categories in Supplementary table; however, when creating a model, they are quantified according to the level of smoking or drinking.

### Data Preparation

The following patients were excluded: 1) patients in whom a variant was found, 2) those who had already developed severe disease at the stage of hospitalisation, 3) those who had undergone hospital transfer, 4) those for whom the day of hospitalisation is unknown and 5) those with hospital-acquired infection and in whom onset occurred ≥15 days prior hospitalisation. Furthermore, it has been reported that age is an important risk factor of severe progression; therefore, a cohort analysis according to age was performed for those aged 40–59, 60–79 and ≥80 years. There were few patients aged < 40 who developed severe disease during hospitalisation; therefore, to ensure the feasibility of the analysis, these patients were not included in our analysis.

### Definitions of severity on admission

When either of the following was required to be provided as respiratory support during hospitalisation, the disease was defined as severe: 1) oxygen therapy (High Flow Nasal Cannula), 2) non-invasive mechanical ventilation (BIPAP and CPAP), 3) invasive mechanical ventilation and 4) extracorporeal membrane oxygenation (ECMO).

### Ethics

This study was approved by the National Center for Global Health and Medicine (NCGM) ethics review (NCGM-G-004133-00). Information regarding opting out of our study is available on the registry website.

### Statistical Analysis

Machine learning was used to predict whether or not progression to severe disease would occur. For constructing the machine learning model, we used the DataRobot enterprise AI platform (DataRobot Automated Machine Learning version 6.3; DataRobot Inc.). For each cohort, the prediction for progression to severe disease was evaluated. Supplementary table shows the positive and negative sample sizes for each cohort. Following the best practice established in the field of machine learning, 20% holdout data was set, and using the remaining data, we conducted five-fold cross-validation, selected the model with the best accuracy and performed prediction for the holdout data. Because the sample size was not large, the 20% holdout method was tested 100 times, and with its mean we tested accuracy. The detailed steps are described below.

1. 20% of the dataset was randomly selected as the holdout and excluded from training in each cohort, as it was suggested that a minimum of 100 events and 100 non-events are required for external validation of prediction models [12,13]. Following the best practice established in the field of machine learning, 5-fold cross validation (CV) was applied to the remaining data to identify the best models [14]. To identify the highest CV, various algorithms were considered including Random forest, Gradient boosted trees, Extreme gradient boosted trees, Light-gradient boosted machine, Neural networks, Regularized regression, Elastic-net, K-Nearest neighbors, Support vector machine, Generalized additive model. For each model, appropriate preprocessing methods were applied. For example, for missing data and, in particular, for linear models, processing was performed such as the creation of a new missing data flag such as with −9999 and interpolation of the median of each algorithm. The hyper-parameters were optimized within each fold by creating an additional fold. Each combination of hyperparameters was tested within the fold to determine optimal hyperparameters. The algorithm was then retrained using these hyperparameters.
2. Using a model with good log-loss accuracy in the cross-validation, we examined multiple ensemble models and searched for the model with the best accuracy. In the ensemble model, we examined the average, median, elastic-net model and generalised linear model and confirmed that three to five models constituted the ensemble.
3. For selecting features, we used permutation importance, which was known as an indicator of how important each feature was in terms of a model’s performance, and it was applicable in any kind of ML model [15]. For the ensemble model with the best accuracy on cross-validation, we created feature sets for the top 30, 40, 50 and 60 features of permutation importance and performed relearning. Permutation feature importance was selected from the top features, and upon deliberating with clinicians, we chose to always include sex, age, no comorbidity, body temperature, white blood cell count and C-reactive protein (CRP) level in the model. We selected the combination of features with the best accuracy, and after relearning with the use of the entire data, except the holdout data, we predicted the holdout. To prevent overfitting, the holdout data was never used other than for verification nor was it used at the stage of narrowing-down the number of features.
4. Random seeds to determine partitioning were changed a hundred times, and the above steps were repeated.
5. Finally, assuming actual application and considering the time-series, we created a model using data recorded before February 2021 and evaluated the data recorded from February 2021 onwards as holdout data, which accounted for approximately 20% of the data recorded during the entire analysis period for all cohorts.

After performance validation, permutation importance computations across 100 simulations were conducted. The computation was applied to the model used for predicting holdout data in each iteration. The permutation importance computations were visualized with the box plot, and we focused on top 20 features in terms of the median. In order to understand the relationship with the severity and each feature, we used partial dependence [16]. Partial dependence plot shows how the average predicted value changes as specified features vary over their marginal distribution. We calculated partial dependence plots for top 5 features in terms of the median of permutation importance.

## Results

Patient data were collected for the period of onset from November 14, 2020 to April 11, 2021. To reduce the amount of variation in the treatment policies of each institution, we used the data of patients with onset after the date shown in the medical treatment guide for patients with severe disease (COVID-19 treatment guidelines, version 5.1) released by the New Coronavirus Infectious Disease Control Promotion Headquarters of the Ministry of Health, Labour and Welfare. We used the data of patients with onset prior to April 11, 2021, when vaccination for elderly individuals belonging to the general population had commenced. On excluding patients who met the exclusion criteria, 14,163 patients were included in our analysis. There were few patients aged < 39 years with severe disease; therefore, such patients were excluded so that the accuracy of risk factor measurements performed using machine learning could be ensured. Patients aged 50–59, 60–79 and ≥80 were divided into three age groups and examined.

Basic attributes of the patients revealed that in all age groups, there were more men, and as age increased, the proportion of women in each group increased. With regard to symptoms, there was no change in body temperature at admission and the proportion of patients with fever according to age. The incidence of coughing, nasal drip and sore throat decreased with age. The feeling of fatigue as well as heart rate decreased in those over 80 years of age.

In terms of patient background, the number of patients with at least one underlying disease, COPD, renal dysfunction and peripheral vascular disorder increased with age. Body mass index and obesity decreased with age; however, there was a higher rate of severe cases among young patients. The incidence of diabetes and hyperlipidemia was high up to 60–79 years of age, which then decreased after 80 years of age. The incidence of hypertension also decreased with age.

Test findings revealed elevated CRP levels in patients aged ≥ 50–69. Albumin levels decreased with age, and LDH levels were high up to 60–79 years of age, which then decreased after 80 years of age. The incidence of pneumonia detected using X-ray and computed tomography increased with age.

The Area Under Curve (AUC) of Receiver operating characteristic (ROC) curve and the area under the precision-recall curve (AUPRC) for each cohort, with the mean specificity, mean sensitivity and 95% CI are presented in Table 1. The numbers presented in brackets indicate the 95% CI. For the CI, we checked the distribution of 100 simulations and used the value closest to the 95% interval. In terms of model accuracy, the 40–59-age group had an AUC of 0.84 (95% CI, 0.78-0.90), sensitivity of 57 % (95% CI, 44%-73%) when we set 90% of specificity; the 60–79-age group had an AUC of 0.82 (95% CI, 0.77-0.87), sensitivity of 52% (95% CI, 43%-65%) and the ≥80-age group had an AUC of 0.76 (95% CI, 0.69-0.81), sensitivity of 41% (95% CI, 28%-54%).

**Table 1.** Indicator of model accuracy.

| Age group | Method of examination | AUC | AUPRC | Specificity = 0.90 | Sensitivity = 0.90 |
| --- | --- | --- | --- | --- | --- |
|  |  |  |  | Sensitivity | Specificity |
| 40-59 | 100 times simulation | 0.84 [0.78, 0.90] | 0.29 [0.17, 0.46] | 0.57 [0.44, 0.73] | 0.58 [0.48, 0.67] |
|  | Time-series examination | 0.87 | 0.26 | 0.62 | 0.56 |
| 60-79 | 100 times simulation | 0.82 [0.77,0.87] | 0.40 [0.32,0.49] | 0.52 [0.43,0.65] | 0.50 [0.42,0.57] |
|  | Time-series examination | 0.82 | 0.42 | 0.49 | 0.51 |
| 80- | 100 times simulation | 0.76 [0.69,0.81] | 0.25 [0.15,0.36] | 0.41 [0.28,0.54] | 0.39 [0.28,0.48] |
|  | Time-series examination | 0.77 | 0.20 | 0.27 | 0.43 |

AUC: Area under curve; AUPRC: Area under precision-recall curve

For each cohort, a boxplot of permutation importance that was obtained by changing the random seed 100 times is shown in Figure 3. As a symptom-related item, respiratory rate was associated with progression to severe disease in younger individuals. In addition, SpO_2_ was the most important factor that was common in those aged ≥60 years. There were more test values than symptoms, and they were included in the risk factors. The presence of an underlying disease was not included as a risk factor. LDH level was a common important factor in all age groups.

**Figure 1.**
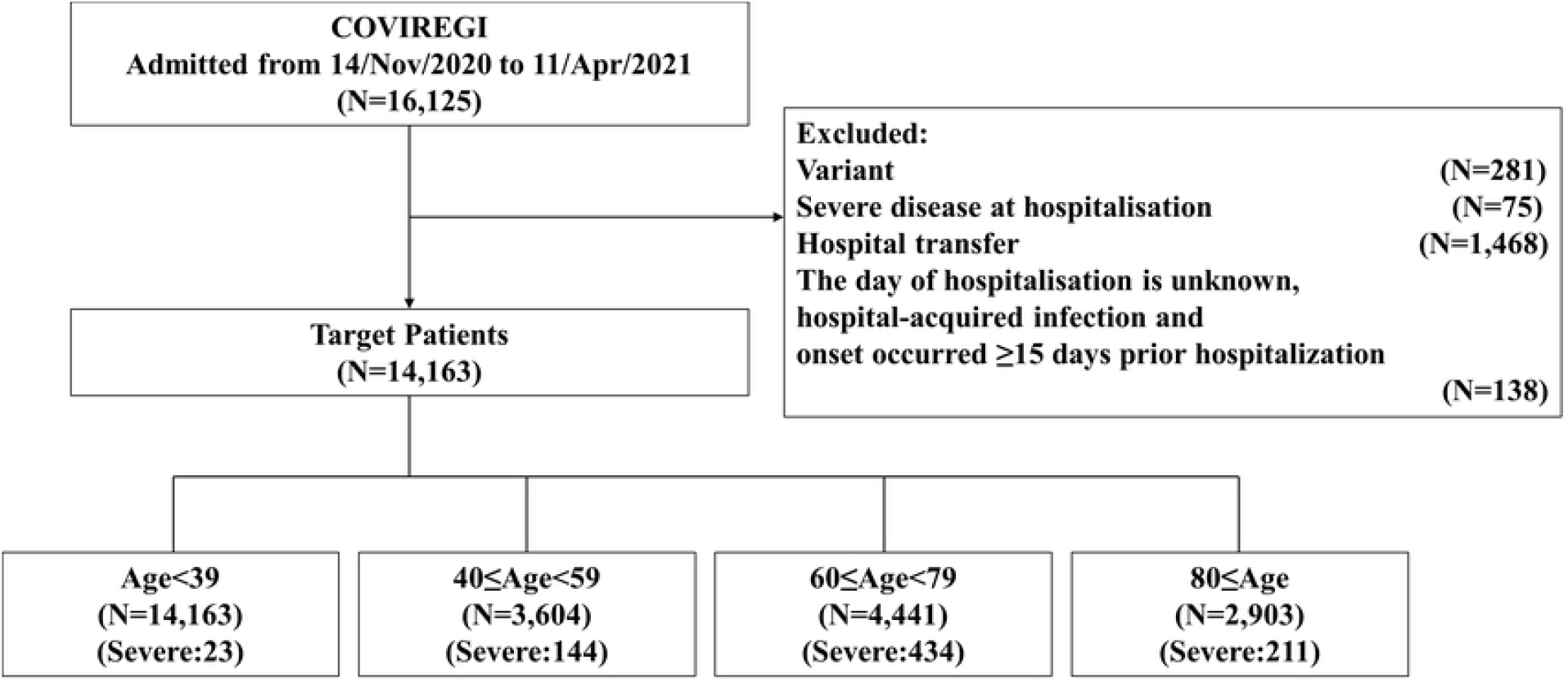
The flowchart of the number of cases. Severe: patients who were required to be provided as respiratory support during hospitalization defined as following, 1) oxygen therapy (High Flow Nasal Cannula), 2) non-invasive mechanical ventilation (BIPAP and CPAP), 3) invasive mechanical ventilation and 4) extracorporeal membrane oxygenation (ECMO)

**Figure 2.**
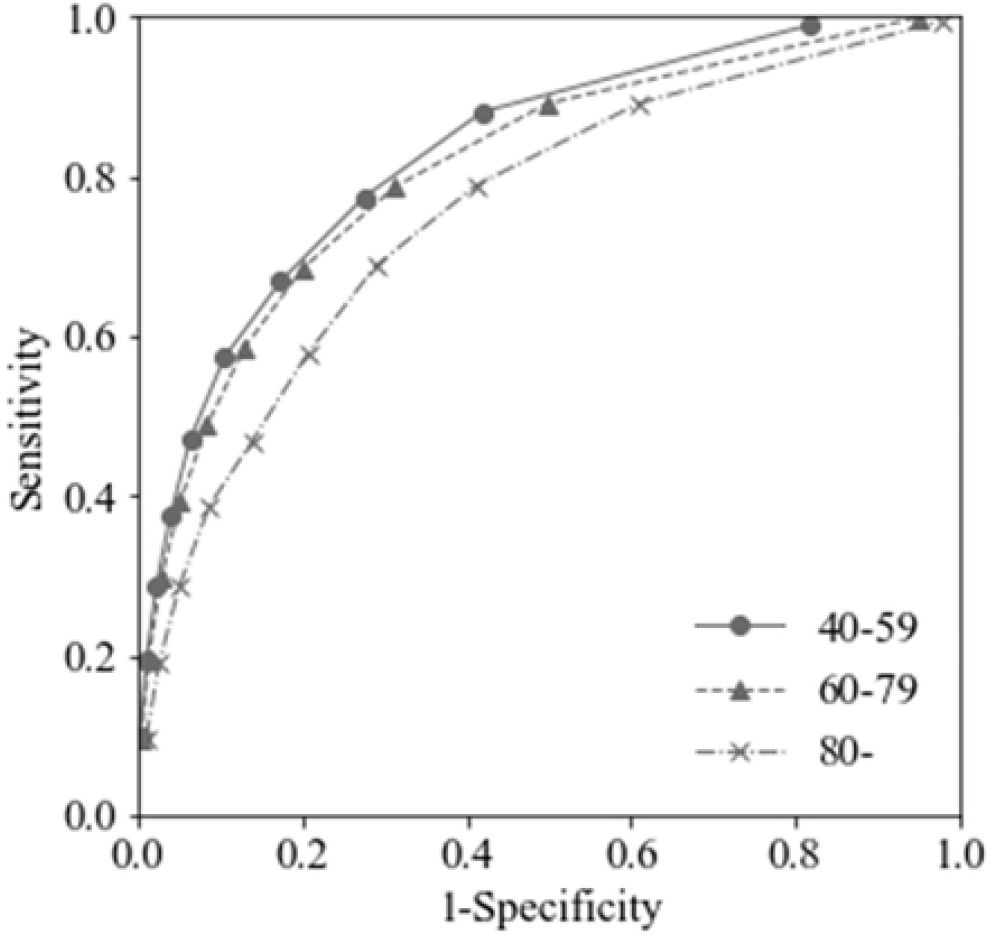
Receiver operating characteristic (ROC) curve of each cohort.

**Figure 3:**
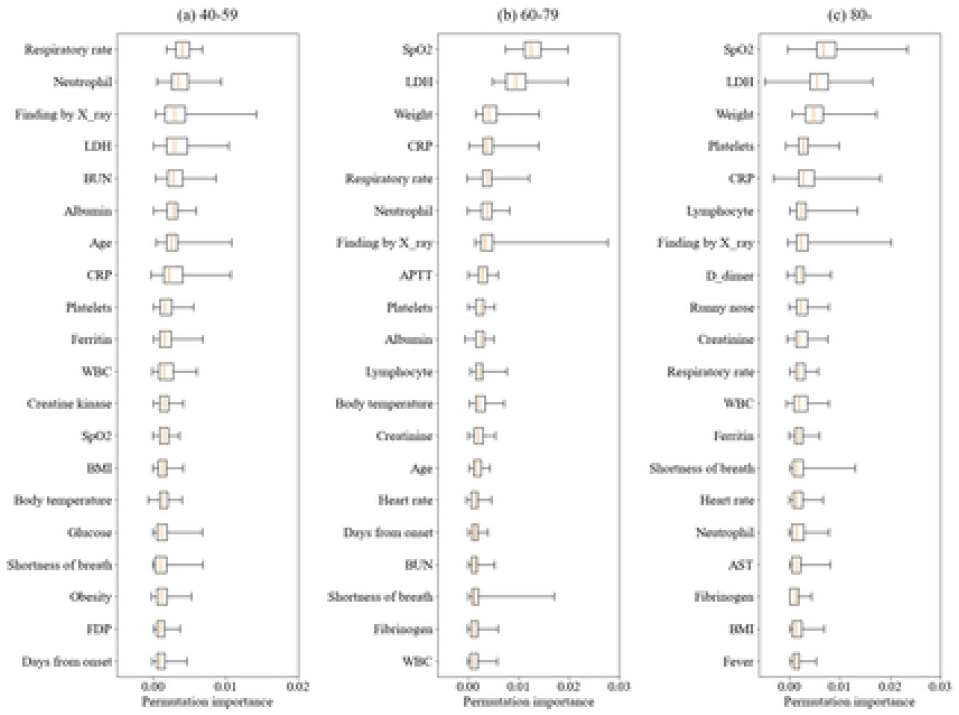
Permutation importance of top features in each cohort.

For the top-ranking item of each feature, the directionality of the effect was confirmed (Figure 4.). Higher respiratory rates and lower SpO2 values corresponded to higher risks of progression to severe disease. In the elderly aged ≥ 80, high body weight posed a risk for progression to severe disease. As CRP and blood urea nitrogen levels increased, the risk or progression to severe disease increased. There was no discrepancy with the observations in clinical settings. Furthermore, the fact that the risk increased as LDH levels increased is considered to be a new finding.

**Figure 4:**
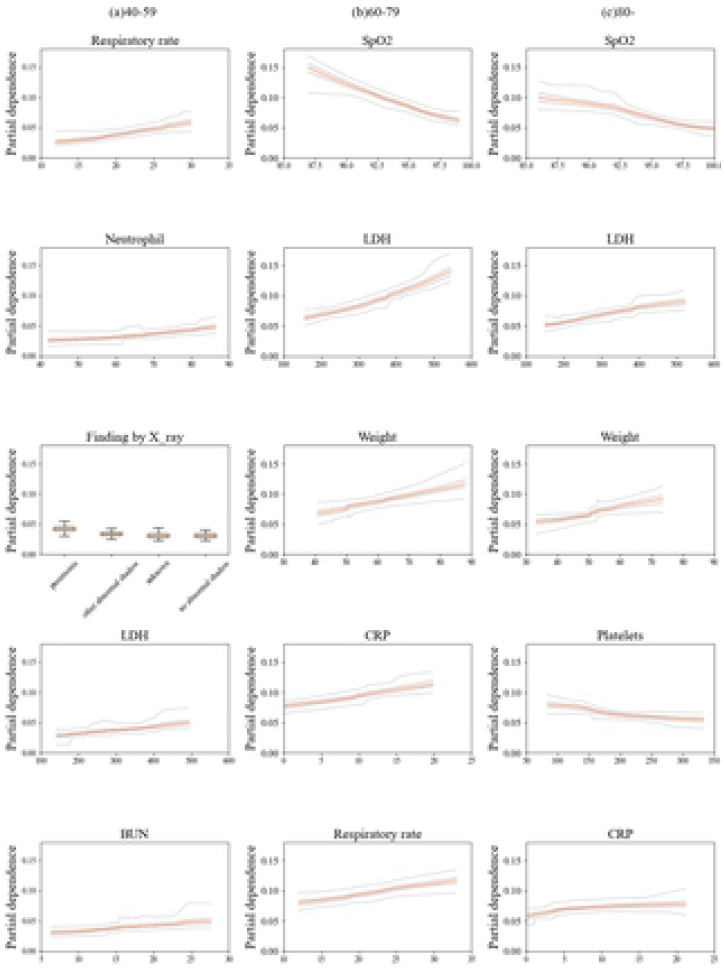
Partial dependence of top features in each cohort. *The results of each feature’s partial dependence: Partial dependence plots of the categorical features across 100 simulations are visualized with box plots, whereas those of numeric features are described with five lines in the order of the maximum value, third quartile, median, first quartile, and minimum value. Orange lines indicate the median.

## Discussion

In the present study, we examined the risk of requiring nasal high flow or higher in patients with COVID-19 using machine learning. Studies conducted to date have mostly targeted death and invasive artificial respiratory management. In Japan, where at-home oxygen facilities and oxygen stations have been established, the efficiency of health resource distribution needs to be improved; therefore, this study targeted patients who ultimately required nasal high flow or higher, which require inpatient management. The AUC, AUCPR, sensitivity and specificity had higher accuracy than the model used for assessing risk scores in the Japanese treatment guideline and were therefore considered to be quite satisfactory results [5,17]. The use of machine learning meant that features could be used in the statistical model without limiting their number, which helped to improve the accuracy [18].

Among individuals aged 40–59, it is necessary to use a highly sensitive index during normal times as well as during medical crisis. If high specificity could pick up approximately 90%, it would enable risk stratification, reducing the number of unnecessary hospitalisations. In individuals aged ≥ 60, there is a high risk of death [19,20]; thus, a highly sensitive index is usually used. When at-home oxygen therapy is desired, careful follow-up can be performed if high specificity is not applicable.

With regard to risk factor details, there is a possibility that the risk may be high in the elderly owing to frailty, and as a result, it was observed that the higher was the body weight, the higher was the risk [21,22]. It has been suggested that the low risk of mortality in Japanese people is associated with lifestyle habits such as obesity [7]. SpO_2_ was not found to be a top-ranking feature in the young generation. In them, it is possible to adequately observe progressions to severe disease on the basis of symptoms alone; therefore, this is a valuable finding, which indicates that SpO2 monitors that are lent in Japan should be prioritised for elderly individuals. Furthermore, because the test values served as a greater risk factor than symptoms, in situations where a patient visits a hospital, top priority should be given to each patient’s condition and predicting the severity of the disease. Additionally, there is an urgent need to develop a system for obtaining detailed information via blood collection.

Furthermore, in this matter, LDH was found to be the top-ranking risk factor. LDH is a parameter for which results can be obtained rapidly; therefore, we believe that it is a useful item for risk estimation in laboratory diagnosis. On the basis of these results, we plan to develop an application and obtain highly accurate risk scores.

The present study had the following limitations. First, the effect of vaccination and variants were not taken into consideration. We plan to update the verification of accuracy, such as the positive predictive value, on the basis of new data in the future. However, it is considered that the priority of the items does not change because the change in severity does not have a great influence on the pathological condition. In this study, we gave priority to rapidity so that it can be used in the next epidemic. Second, the registry is not inclusive; it is a registry that targets inpatients. In Japan, there were changes to the hospitalisation criteria as well [4]. However, in the present study, we limited the time period and tried to organise the hospitalisation criteria as much as possible. Furthermore, in Japan, there are some hospitals with >200 beds and patients with moderate–severe illness, and because the hospitals are equipped with sufficient facilities, we believe that there is no major difference in practice. Third, there is no guideline or criteria for using nasal high flow; therefore, it partially depends on the medical resources of the institution and the practice of each physician. For this reason, accuracy was underestimated.

## Conclusion

We were able to estimate the risk of requiring nasal high flow or higher in patients with COVID-19 using machine learning. On the basis of these results, we plan to develop an application for obtaining highly accurate risk scores, which can be used in clinical settings.

## Data Availability

The datasets generated during and/or analyzed during the current study are not publicly available due to the regulation of COVIREGI-JP. A portion of the data would be available upon reasonable request to the corresponding author.

## Acknowledgments

We thank all the participating facilities for their care of COVID-19 patients, and cooperation on data entry to the registry.

## Author contributions

NM conceived the study. KK, YS, and SI developed and validated the model. YA, ST, TA, JY, and GY interpreted the results. YA, ST, TA, JY, MT, SetsukoS, and KS curated the data. NM and ST wrote the draft of the manuscript. NM and NO lead the project. All the authors critically reviewed the manuscript and approved the final version.

